# Telerehabilitation services have declined post-COVID-19

**DOI:** 10.1101/2024.07.22.24310787

**Authors:** Anthony K May, Anne E Holland, Jennifer A Alison, Kelcie Herrmann, Narelle S Cox

## Abstract

**Objective:** To characterize Pulmonary rehabilitation (PR) service delivery, investigate the impact of the pandemic on PR services, and describe centre-based PR (CBPR) and telerehabilitation with reference to PR essential components.

**Design:** Online national cross-sectional survey.

**Setting:** Australian PR services.

**Participants:** Representatives of PR programs listed within the Lung Foundation Australia national database (n=295).

**Interventions:** Not applicable.

**Main Outcome Measure(s):** Availability of PR in CBPR and telerehabilitation settings.

**Results:** 97% of Australian PR services (n=114/117) delivered CBPR, similarly to pre-COVID-19 pandemic availability (96%). 43% (n=50/116) of services delivered telerehabilitation, which was significantly less than availability during COVID-19 restrictions (74%; p<0.001). CBPR was primarily delivered in a group setting (99%; median (IQR) 7 (6-8) participants/group), and telerehabilitation primarily via individual telephone calls (94%). 39% of respondents report CBPR group size has reduced. PR essential components of initial centre-based assessments and individually prescribed/progressed endurance and resistance training were achieved by most CBPR and telerehabilitation programs. Staff training in delivery of telerehabilitation models was undertaken in 33% of services.

**Conclusions:** PR essential components are generally met in current Australian programs. However, telerehabilitation services and CBPR program capacity have declined indicating reduced program capacity. Sustainability of effective PR programs is required to support access for people with chronic respiratory diseases.

## Introduction

Pulmonary rehabilitation (PR) is a highly effective, yet widely underused treatment for people with chronic respiratory disease [1, 2]. Typically, PR is an 8-12 week program delivered within a hospital or healthcare centre [2]. Limited program availability and patient-related barriers to centre-based PR (CBPR) attendance have increased interest in alternative PR models utilising telerehabilitation to improve program access [2, 3, 4]. Telerehabilitation use expanded during the COVID-19 pandemic. However, post-pandemic it is unclear how many services continue to deliver CBPR and telerehabilitation.

Telerehabilitation programs may use synchronous (e.g. telephone calls, video-conferencing) or asynchronous communication (e.g. email) [3], and can be available across a variety of platforms. A recent Cochrane review has demonstrated that telerehabilitation achieves similar clinical outcomes to CBPR with greater program completion rates [4]. For telerehabilitation to be a clinically acceptable alternative to CBPR, models should meet similar standards to CBPR in delivering essential components of effective PR [5]. Defined PR essential components include initial centre-based assessment, individually prescribed/progressed endurance and resistance training, and delivery by healthcare professionals trained in the specific telerehabilitation model [1]. The extent to which telerehabilitation models deliver essential components of PR in clinical practice is not clear.

This study aimed to characterise PR service delivery, investigate the impact of the pandemic on PR services, and describe CBPR and telerehabilitation with reference to PR essential components.

## Methods

An online, cross-sectional survey was undertaken between July 19 and August 28, 2023 (Qualtrics, Provo, UT, USA), with pilot testing prior to launch. Email invitations for voluntary anonymous survey completion were sent to all PR programs within the Lung Foundation Australia national database, the most comprehensive record of programs available. Ethics approval was granted prospectively (Monash University (ID 39264)).

The survey comprised twenty-seven questions (plus sub-questions as required) that explored the study aims. Demographic information relating to respondent role and PR service setting were collected.

All responses received, including from incomplete surveys, were included in data analysis (IBM SPSS Statistics V28.0 (IBM Corp., NY, USA)). Descriptive statistics were reported (number (%) or median (interquartile range (IQR))). Open responses were coded thematically. Service availability at the time of survey completion was compared with availability pre-COVID-19 pandemic for CBPR, and during the pandemic (2020-22) for telerehabilitation (McNemar’s test; significance p<0.05).

## Results

Survey invitations were sent to 295 PR programs with 117 responses received (40% response rate; n=9 (8%) incomplete). 92% of respondents were the service PR coordinator. For PR service availability and respondent demographics see Figure 1.

**Figure 1:**
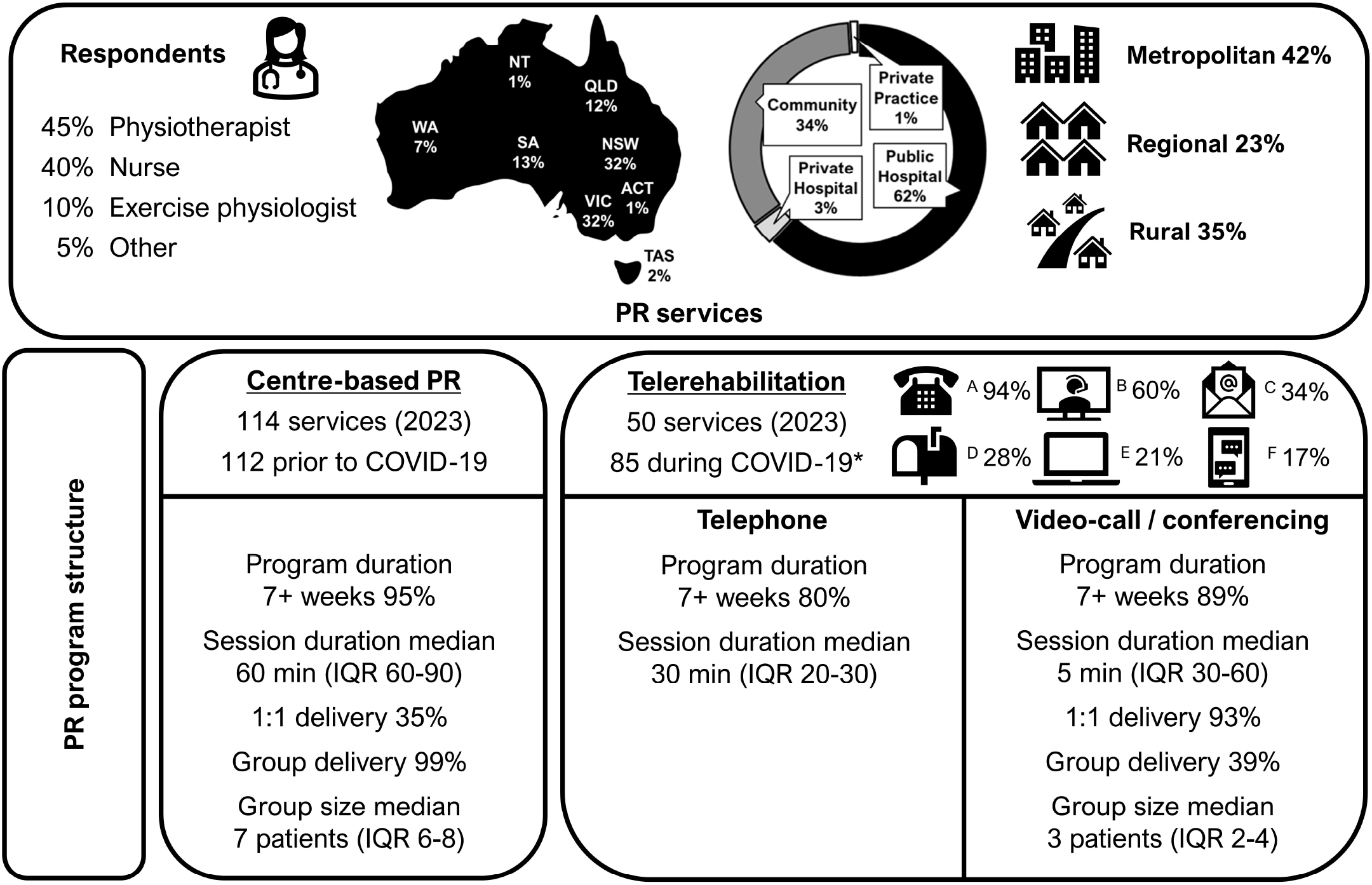
Summary of Australian pulmonary rehabilitation (PR) services and programs. * p<0.001 vs. 2023 (McNemar test). A Telephone; B Video-call/conference; C Email; D Postal service; E Desktop or mobile application; F Text message. ACT Australian Capital Territory; NSW New South Wales; NT Northern Territory; QLD Queensland; SA South Australia; TAS Tasmania; VIC Victoria; WA Western Australia.

97% of respondents (n=114/117) reported delivery of CBPR at survey completion (Figure 1), similar to pre-pandemic CBPR availability (96%). CBPR was primarily delivered in a group setting (n=109/110, 99%), to median (IQR) 7 (6-8) participants/group, which 39% of respondents (n=42/109) reported to be smaller than pre-pandemic group size. The most common CBPR training modalities were walking (90%), free/machine weights (78%), stationery cycling (68%) and resistance bands (53%).

During the pandemic, 74% of respondents (n=85/114) delivered telerehabilitation, which had declined significantly at the time of survey completion (43%, n=50/116; p<0.001) (Figure 1). The most cited reasons for telerehabilitation cessation were staffing limitations, patient preference for CBPR, and staff perception for greater ease/benefits of CBPR. All services except one delivered telerehabilitation in addition to CBPR. Multiple telerehabilitation models were used, including telephone (94%), video-interaction (60%) and email (34%). Of synchronous (video) telerehabilitation programs (n=28), group video-conferencing (n=11/28, 39%; median (IQR) 3 (2-4) participants/session) was less commonly delivered than 1:1 video-calls (n=26/28, 93%). The most common telerehabilitation training modalities were walking (89%), free/machine weights (63%), bodyweight resistance exercises (58%) and resistance bands (50%).

The essential component of initial centre-based assessment was performed in 100% of CBPR and 89% of telerehabilitation programs (Figure 1), while individually prescribed/progressed endurance and resistance training was delivered by most CBPR (91%) but fewer telerehabilitation programs (78%). Staff training in the delivery of specific telerehabilitation models was undertaken in 33% of services (n=15/45).

## Discussion

This study characterises availability and delivery of PR in Australia. CBPR program availability is largely consistent with pre-pandemic levels, but with reduced group size. Meanwhile, telerehabilitation availability has declined compared with during COVID-19 restrictions, although remains higher than pre-pandemic [6, 7]. The majority of telerehabilitation programs complied with PR essential components.

The decline in telerehabilitation availability post-COVID reflects a trend across telemedicine more broadly. Increasing patient preference for in-person consultation, waning concerns about COVID-19 infection and variable administrative and regulatory support for hybrid care delivery models (i.e. face-to-face and telehealth) have all been posed as contributors to reduced telehealth availability [8]. Given that telerehabilitation is a recommended alternative to CBPR in international guidelines [2], and clinical services demonstrated ability to deliver telerehabilitation under COVID conditions, understanding the factors that underpin reduced telerehabilitation provision currently is important if models are to be sustainable.

While most telerehabilitation programs complied with essential components of centre-based assessment and exercise training prescription/progression, relatively few provided telerehabilitation model-specific training. Experience and competency with technology are known factors in the successful delivery of remote healthcare [9]. Whether enhanced telerehabilitation model-specific training would improve clinician confidence and acceptance of telerehabilitation, leading to greater service availability, remains to be determined.

Potential to improve PR service access is a proposed benefit of telerehabilitation models [1, 2]. This study highlighted reduced CBPR group size post-pandemic, along with few telerehabilitation models being delivered in a group format. This creates the very real possibility that overall PR program capacity has actually reduced post-pandemic, further impeding program access for patients. Whether changes in program funding, or other contributors such as referral practices, have contributed to reduced service capacity requires exploration. In Australia, healthcare is largely funded under a universal scheme for subsidisation and reimbursement, however in regions where PR reimbursement is complex, such as the US, fluctuating service availability based on financial drivers may have profound effects on access to PR for patients [5].

The cross-sectional nature of this work relied upon participant recollection of service delivery over the previous 4-year period. This need for recall, coupled with potential for staff changes during the intervening period, may have impacted historical program knowledge held by the respondent. The response rate for this study was 40%. This may be attributed to the online method of survey delivery without incentive [10]. However, given that all Australian states and territories are represented, including rural, regional and metropolitan services, we believe the data to be largely reflective of the current state of Australian PR.

## Conclusions

Most Australian telerehabilitation programs currently meet PR essential components, supporting the ability of such models to deliver effective PR programs. However, telerehabilitation services and CBPR program capacity have both declined post-COVID highlighting the importance of ensuring sustainability of effective PR programs, irrespective of model of delivery, to support widespread access to this recommended treatment.

## Data Availability

All data produced in the present study are available upon reasonable request to the authors

## Acknowledgements

We would like to extend our thanks Emma Halloran for her input during survey development.

## Abbreviations list

CBPR: Centre-based pulmonary rehabilitation
IQR: Interquartile range
PR: Pulmonary rehabilitation

## References

1. Holland AE, Cox NS, Houchen-Wolloff L, et al. Defining modern pulmonary rehabilitation. An official American Thoracic Society Workshop Report. Ann Am Thorac Soc. 2021;18(5):e12–e29.

2. Rochester CL, Alison JA, Carlin B, et al. Pulmonary rehabilitation for adults with chronic respiratory disease: An official American Thoracic Society Clinical Practice Guideline. Am J Respir Crit Care Med. 2023;208(4):e7–e26.

3. Bhatt SP, Rochester CL. Expanding implementation of tele-pulmonary rehabilitation: The new frontier. Ann Am Thorac Soc. 2022;19(1):3–5.

4. Cox NS, Dal Corso S, Hansen H, et al. Telerehabilitation for chronic respiratory disease. Cochrane Database Syst Rev. 2021;1:CD013040(1).

5. Bhatt SP, Casaburi R, Mosher CL, et al. Telehealth pulmonary rehabilitation: A call for minimum standards. Am J Respir Crit Care Med.210(2):156–46.

6. Spruit MA, Pitta F, Garvey C, et al. Differences in content and organisational aspects of pulmonary rehabilitation programmes. Eur Respir J. 2014;43(5):1326–37.

7. Johnston CL, Maxwell LJ, Alison JA. Pulmonary rehabilitation in Australia: A national survey. Physiotherapy. 2011;97(4):284–90.

8. Huang J, Yeung AM, Eiland LA, et al. Telehealth Fatigue: Is It Real? What Should Be Done? J Diabetes Sci Technol. 2024;18(1):196–200.

9. Inskip JA, Lauscher HN, Li LC, et al. Patient and health care professional perspectives on using telehealth to deliver pulmonary rehabilitation. Chron Respir Dis. 2018;15(1):71–80.

10. Shih T, Fan X. Comparing response rates in e-mail and paper surveys: A meta-analysis. Educ Res Rev. 2009;4(1):26–40.

